# International observational survey of the effectiveness of personal protective equipment during endoscopic procedures performed in patients with COVID-19

**DOI:** 10.1101/2020.12.01.20240853

**Authors:** Ryota Niikura, Mitsuhiro Fujishiro, Yousuke Nakai, Koji Matsuda, Takuya Kawahara, Atsuo Yamada, Yosuke Tsuji, Yoku Hayakawa, Kazuhiko Koike

## Abstract

**Introduction and aims:** This international survey was performed to evaluate the cumulative incidence of nosocomial novel coronavirus disease 2019 (COVID-19) among healthcare professionals during endoscopic procedures.

**Methods:** We performed an international web-based self-reported questionnaire survey. Participants completed the questionnaires every week for 12 weeks. The questionnaire elicited responses regarding the development of COVID-19 and details of the PPE used.

**Results:** All 483 participants were included in the analysis. Participants had a mean age of 42.3 years and comprised 68.3% males. The geographic distribution of the study population was Asia (89.2%), Europe (2.9%), North and South America (4.8%), Oceania (0.6%), and Africa (1.5%). The most common endoscopy-related role of the participants was endoscopist (78.7%), and 74.5% had > 10 years of experience. Fourteen participants had performed 83 endoscopic procedures in patients positive for COVID-19. During the mean follow-up period of 4.95 weeks, there were no cases of COVID-19 when treating COVID-19 positive patients. The most common PPE used by participants treating patients with COVID-19 was a surgical mask plus N95 mask plus face shield, goggles, cap, long-sleeved isolation gown, and single pair of gloves. The most common PPE used by participants treating patients without COVID-19 was a surgical mask, no face shield but goggles, cap, long-sleeved isolation gown, and single pair of gloves during all endoscopic procedures.

**Conclusions:** The risk of COVID-19 transmission during any endoscopic procedure was low in clinical practice.

## Introduction

The morbidity and mortality of novel coronavirus disease 2019 (COVID-19) are increasing worldwide. The virus responsible for this disease, severe acute respiratory syndrome coronavirus-2 (SARS-CoV-2), is highly infectious and causes nosocomial infection in healthcare professionals [1]. Especially, endoscopic procedures have a potentially high risk for healthcare professionals due to transmission via aerosol from COVID-19 patients [2-7]. In addition, SARS-CoV-2 has also been detected in stool specimens, suggesting a possible risk of fecal–oral transmission [8], similar to SARS [9,10]. However, although endoscopic procedures pose a risk of COVID-19 transmission, they are often necessary for diagnosis and treatment of gastrointestinal (GI) bleeding, malignancies, obstructive jaundice, and cholangitis. Recently, a research group in Italy [11] and the World Endoscopy Organization [12] performed a hospital-based email questionnaire survey among endoscopy units and clarified the current status of personal protective equipment (PPE) use when performing endoscopic procedures in patients with a positive diagnosis or suspected COVID-19. Although these data represent very important initial reports, we considered the further necessity of data regarding the type of PPE used during endoscopic procedures in patients with different risks of COVID-19 transmission. The incidences of SARS-CoV-2 genomic mutations and clinical COVID-19 reportedly vary among geographic regions [13]. Therefore, we designed an international web-based questionnaire survey for endoscopy-related healthcare professionals (UMIN000040162) consisting of 16 questions regarding the use of PPE, such as surgical masks, face shields, goggles, gowns, and gloves. The aims of this survey were to evaluate cumulative short-term incidence rates of COVID-19 and to share information among healthcare providers regarding the appropriate PPE use to prevent COVID-19 transmission.

## Methods

### Design and participants

We performed an international web-based questionnaire survey among endoscopy-related healthcare professionals, including endoscopists, nurses, laboratory technicians, and endoscopy reprocessing technicians working in the endoscopy suite. Participants were invited to join the survey by the principal investigators related to personal communication and World Endoscopy Organization volunteer members and were enrolled by snow-ball sampling methods. The survey began on April 15, 2020 and ended on August 8, 2020. Participants completed web-based questionnaires every week for 12 weeks, prompted by automated reminder emails using Google Forms®. The study was approved by the Ethics Committee of the University of Tokyo Hospital (No. 2020046NI).

### Web-based questionnaire

The web-based questionnaire included items regarding COVID-19 development, age, sex, role (endoscopist, nurse, laboratory technician, endoscopy reprocessing technician), geographical area (North America, South America, Africa, Asia, Europe, Oceania), number of years of experience, PPE (mask and face shield, goggles and face shield, cap, gown, gloves), endoscopic procedures (esophagogastroduodenoscopy (EGD), colonoscopy, endoscopic retrograde cholangiopancreatography (ERCP), endoscopic ultrasound (EUS), endoscopic hemostasis, endoscopic submucosal dissection (ESD)) performed in patients with and without confirmed COVID-19, and PPE replacement frequency (After each procedure, every day, 1-7day, every week). N95 masks were defined as N95 (United States NIOSH-42CFR84), FFP2 (Europe EN 149-2001), KN95 (China GB2626-2006), P2 (Australia/New Zealand AS/NZA 1716:2012), Korea 1st class (Korea KMOEL - 2017-64), and DS (Japan JMHLW-Notification 214, 2018) masks. The details of the questionnaire are shown in Supplementary Table 1.

**Table 1.**
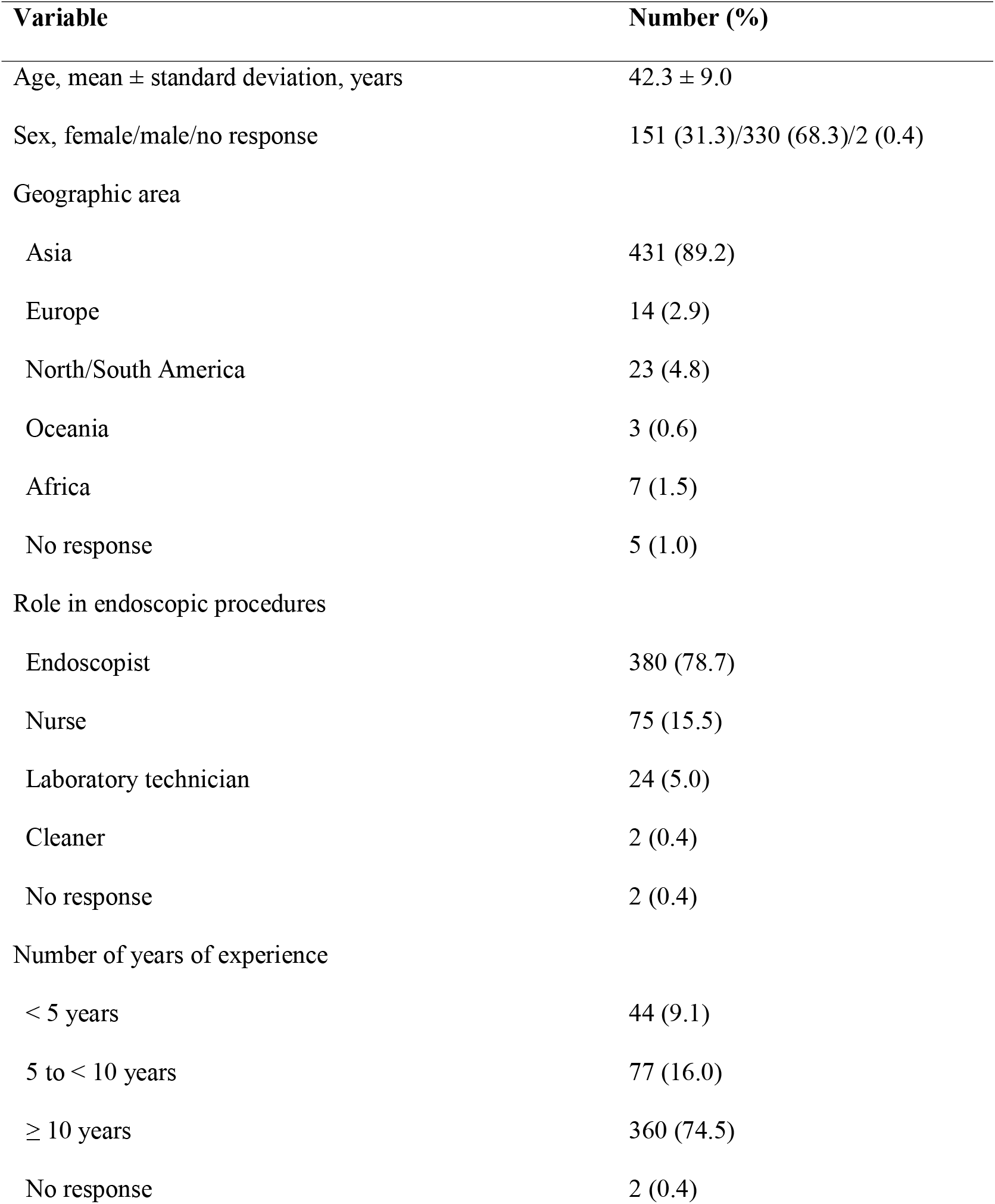
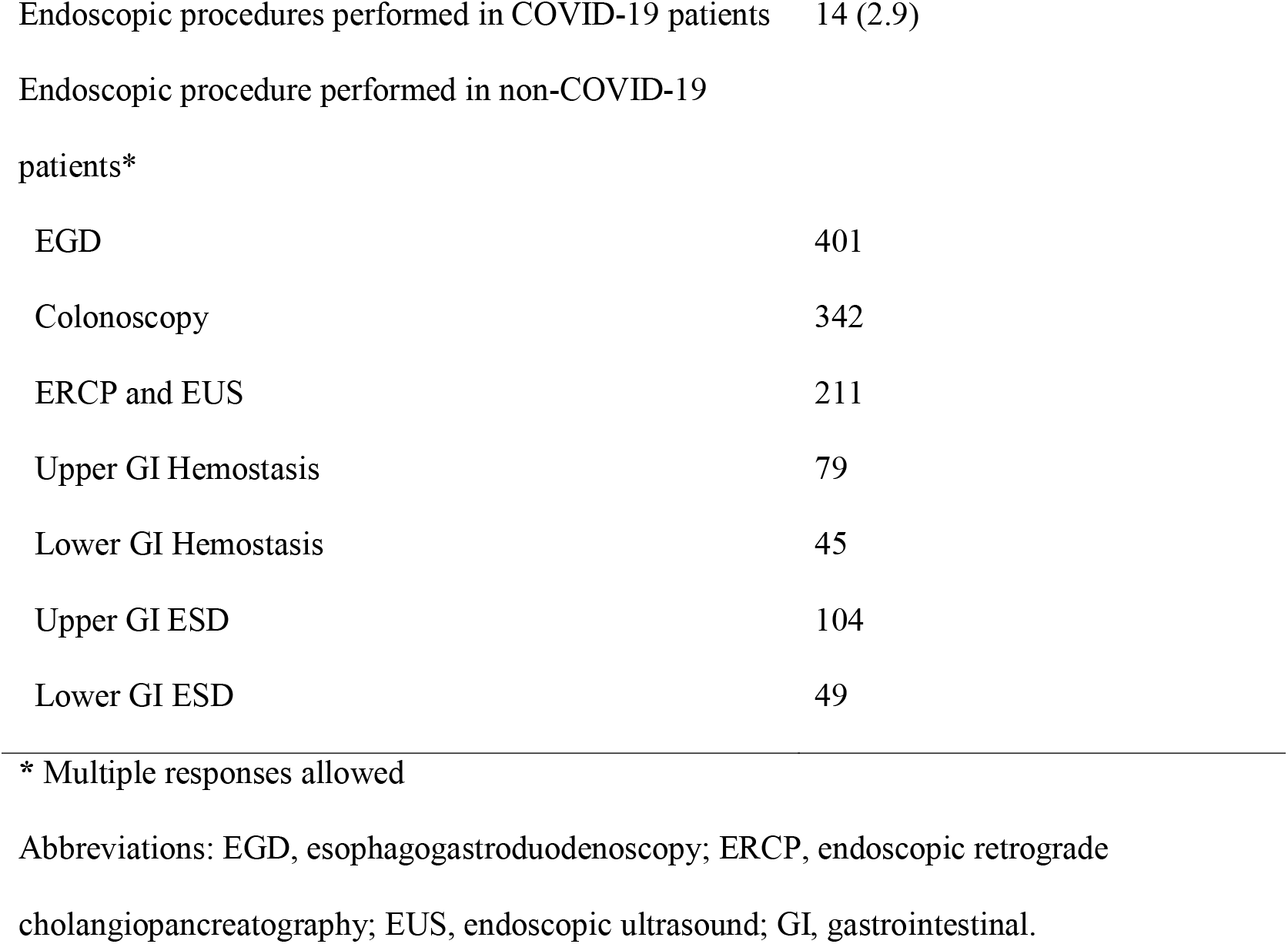
Participant characteristics (*n* = 483)

### PPE evaluation

We evaluated each PPE for protection of the mouth, eyes, body, and hands and defined each as mask and face shield, goggles and face shield, cap and gown, and gloves. We also defined the minimum and maximum use of each PPE category as follows. For the mask and face shield category, the minimum PPE use consisted of a surgical mask, followed by N95 mask, and the maximum was N95 mask plus face shield. Regarding goggles and face shields, the minimum was no goggles or face shield, followed by face shield but no goggles, and goggles but no face shield; the maximum PPE was use of both goggles and face shield. For caps, the minimum and maximum were no use and use of a cap, respectively. Regarding gowns, the minimum was short-sleeved vinyl-type gown, followed by short-sleeved vinyl-type gown plus arm covers, and long-sleeved vinyl-type gown; the maximum was long-sleeved isolation-type gown. For gloves, the minimum and maximum uses were a single pair and double pair of gloves, respectively.

### Statistics

The full analysis set included participants who had not been diagnosed with COVID-19 at baseline to exclude potential cases of community-acquired or nosocomial infection not related to endoscopic procedures. The primary outcome was the incidence of COVID-19 when treating COVID-19 positive patients, defined as a self-reported positive PCR test in the absence of close contact with an infected subject other than patients among participants who performed endoscopic procedures for COVID-19-positive patients; this was censored at the date of completion of the final questionnaire. The secondary outcome was the incidence of COVID-19 when treating patients without confirmed COVID-19.

The Kaplan–Meier method was used to estimate the cumulative incidence of COVID-19 over a 12-week period from the start to questionnaire response was calculated. Number of COVID-19 new cases were evaluated using public site data (https://github.com/owid/covid-19-data/tree/master/public/data). The statistical analyses were performed using SAS software (version 9.4; SAS Institute, Cary, NC).

## Results

From April 15 to August 8, 2020, a total of 488 participants were invited to join the survey. After excluding 5 because of a diagnosis of COVID-19 at baseline, 483 participants were included in the analysis. Of 483 participants, 103 participants (21.3%) completed the questionnaire during the 12 weeks of the study. Number of COVID-19 new cases during study period is shown in **Figure 1**.

**Figure 1.**
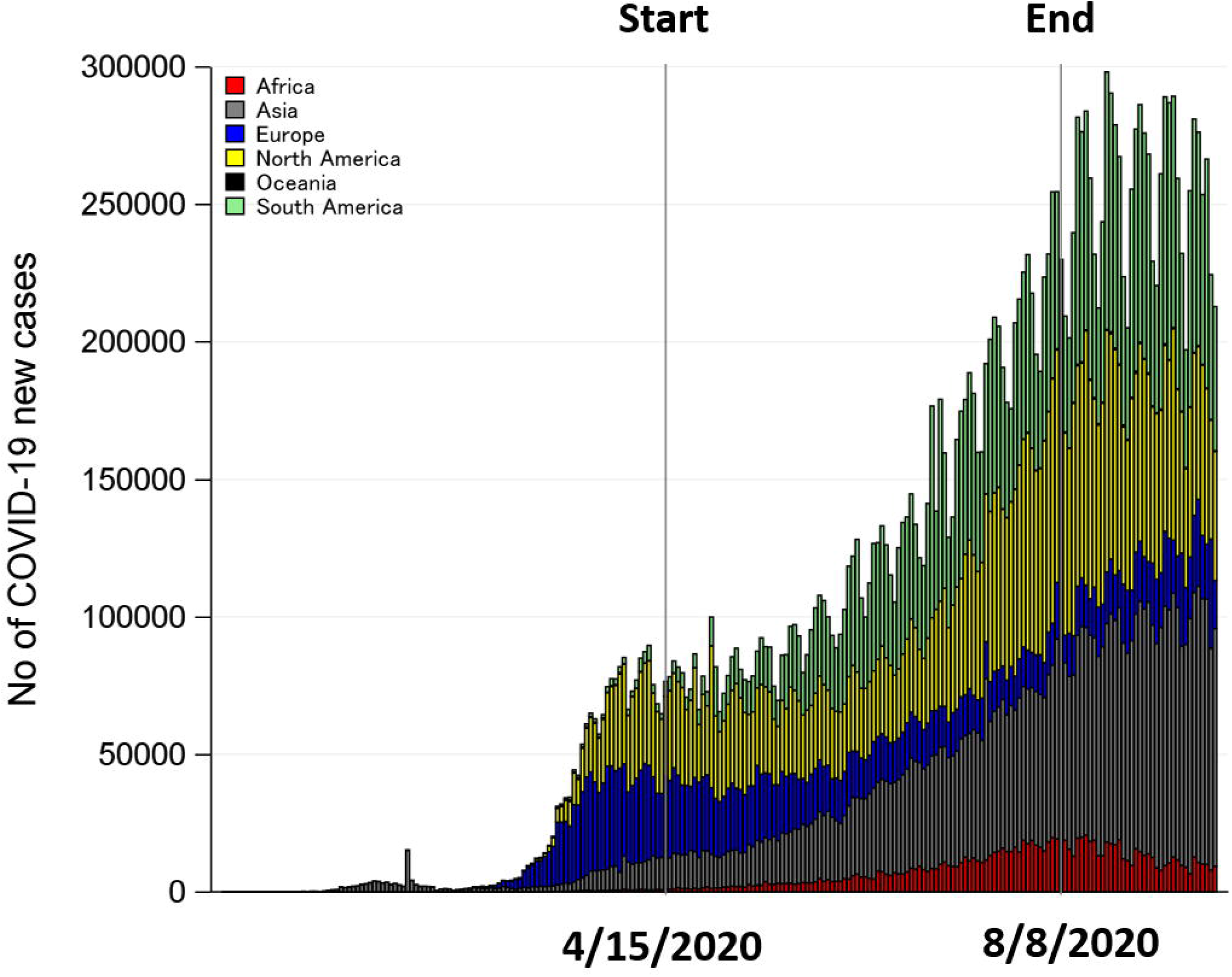
COVID-19 pandemic during study period

### Participant characteristics and endoscopic procedures

The participants had a mean age of 42.3 years and comprised 68.3% (330/483) males. The distribution according to geographic area was as follows: Asia (89.2%), Europe (2.9%), North and South America (4.8%), Oceania (0.6%), and Africa (1.5%). The most common endoscopy-related role of the participants was endoscopist (78.7%), and 74.5% had more than 10 years of experience. The participants had performed EGD (401), colonoscopy (342), ERCP and EUS (211), upper GI hemostasis (79), lower GI hemostasis (45), upper GI ESD (104), and lower GI ESD (49; multiple responses included). Change of each endoscopic procedure volume during study period are shown in **Figure 2**. EGD volume was increased (A), and other procedure volumes were same level at beginning and end of the study (B-E).

**Figure 2.**
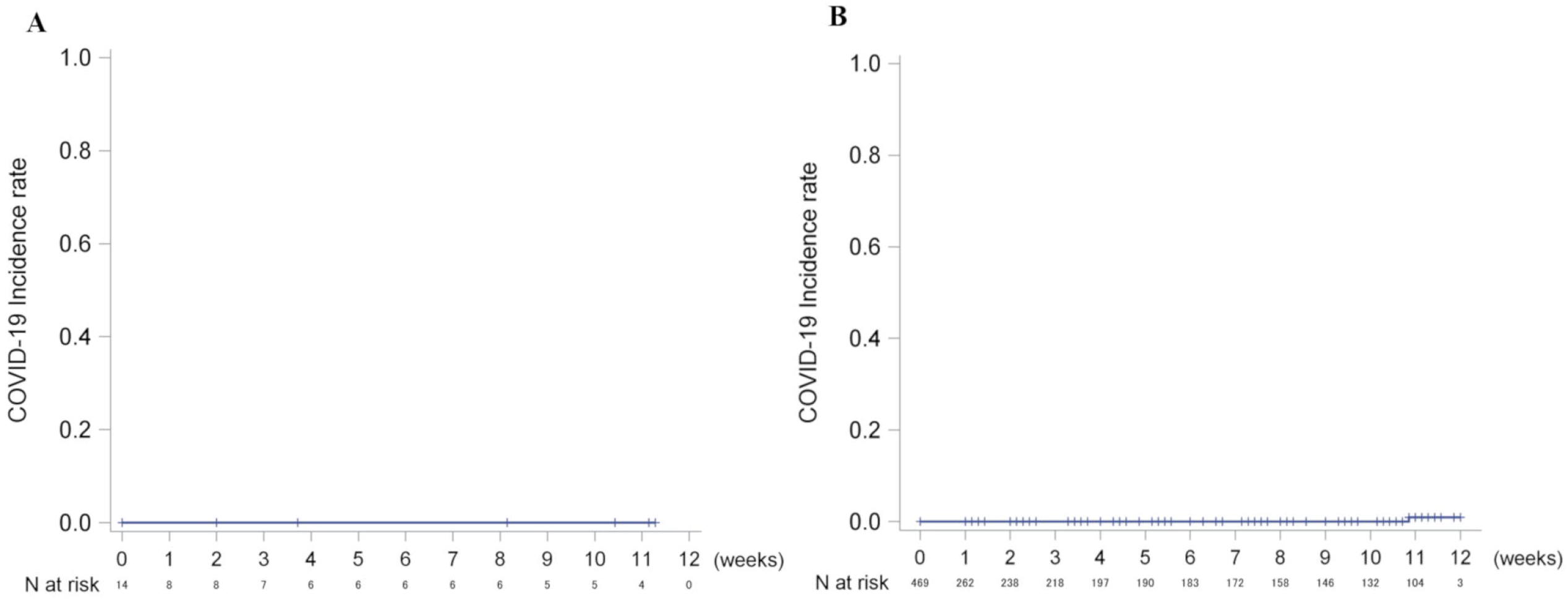
Change of endoscopic procedure volume during study period (A) 10 or more upper endoscopy (B) 10 or more colonoscopy (C) One or more ERCP and EUS (D) One or more ESD (E) One or more upper and lower GI hemostasis Bar shows 95% confidence interval.

### COVID-19 incidence and the minimum and most common PPE used when treating COVID-19-positive patients

Fourteen participants performed 83 endoscopic procedures in patients with a positive diagnosis of COVID-19. None of these participants developed COVID-19 over the mean follow-up period of 4.95 weeks (**Figure 3A**). The distribution according to geographic area was as follows: Asia, 43%; Europe, 22%; North and South America, 21%; Oceania, 7%; and Africa, 7%. The procedures performed were EGD by 11 participants, colonoscopy by 7, ERCP and EUS by 10, hemostasis during EGD by 6, hemostasis during colonoscopy by 3, and upper GI ESD by 2. The details are shown in **Table 2**.

**Table 2.**
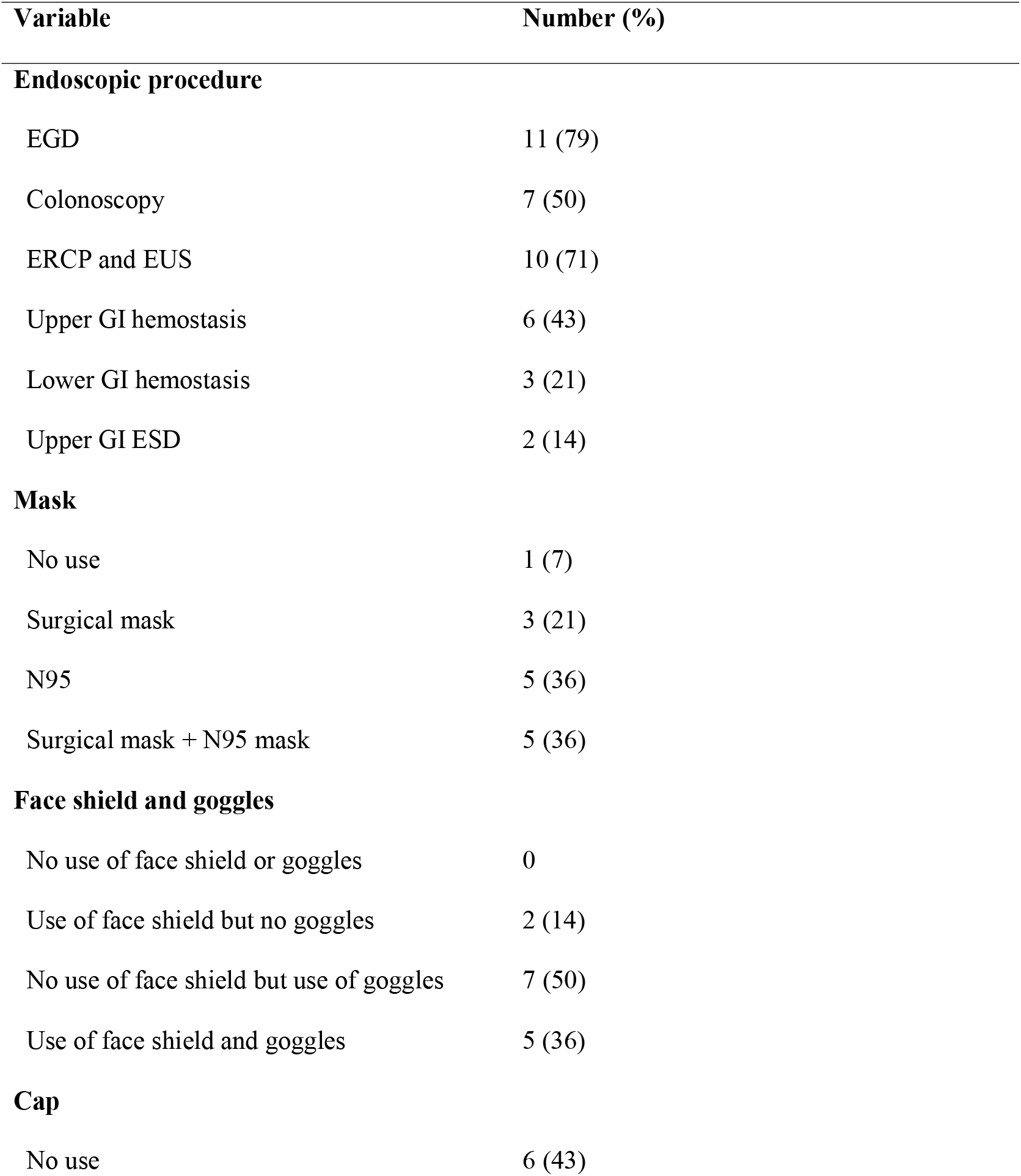
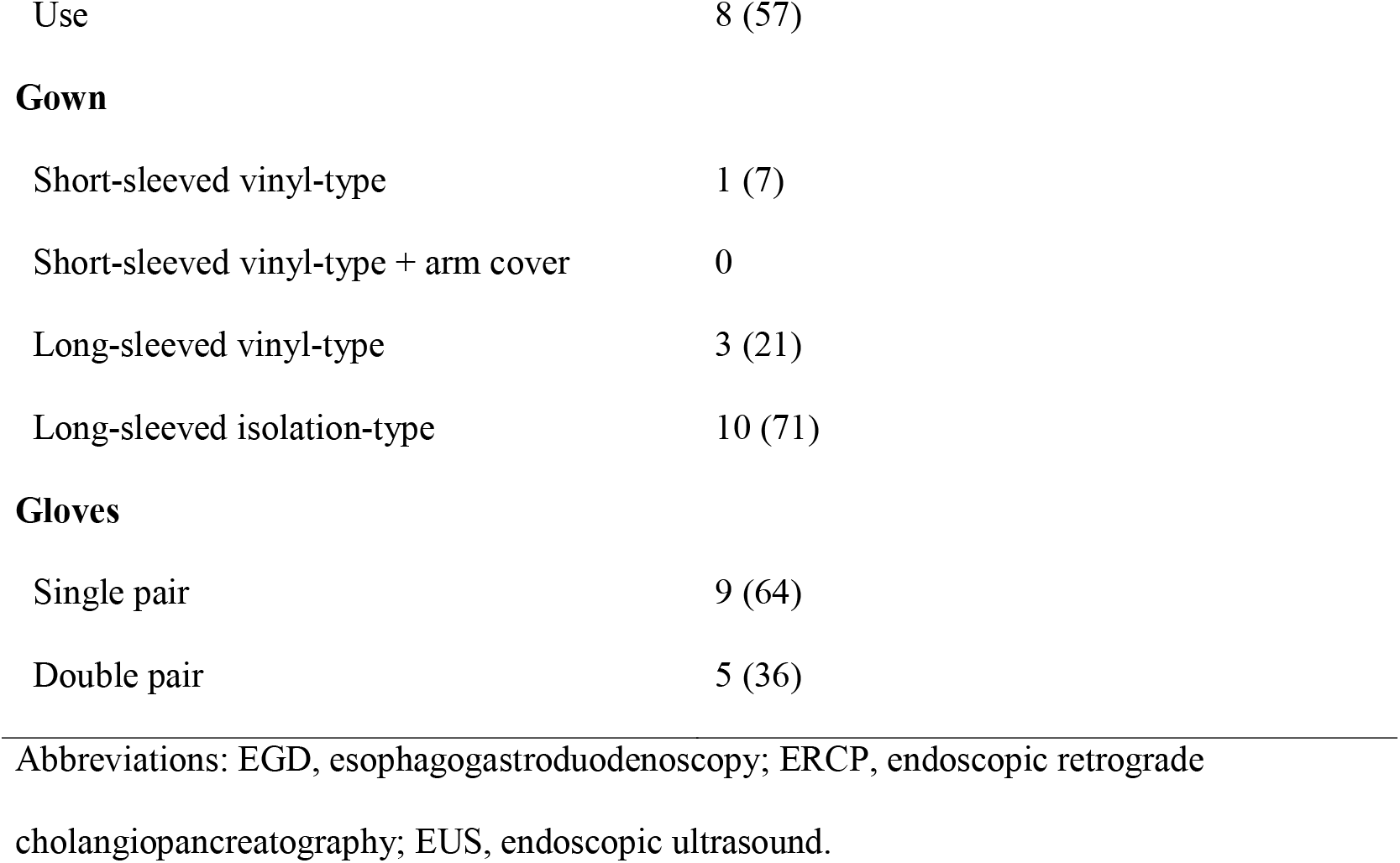
Endoscopic procedures and personal protective equipment used during procedures performed in patients with confirmed COVID-19 (*n* = 14)

| Variable | Number (%) |
| --- | --- |
| **Endoscopic procedure** |  |
| EGD | 11 (79) |
| Colonoscopy | 7 (50) |
| ERCP and EUS | 10 (71) |
| Upper GI hemostasis | 6 (43) |
| Lower GI hemostasis | 3 (21) |
| Upper GI ESD | 2 (14) |
| **Mask** |  |
| No use | 1 (7) |
| Surgical mask | 3 (21) |
| N95 | 5 (36) |
| Surgical mask + N95 mask | 5 (36) |
| **Face shield and goggles** |  |
| No use of face shield or goggles | 0 |
| Use of face shield but no goggles | 2 (14) |
| No use of face shield but use of goggles | 7 (50) |
| Use of face shield and goggles | 5 (36) |
| **Cap** |  |
| No use | 6 (43) |
| Use | 8 (57) |
| **Gown** |  |
| Short-sleeved vinyl-type | 1 (7) |
| Short-sleeved vinyl-type + arm cover | 0 |
| Long-sleeved vinyl-type | 3 (21) |
| Long-sleeved isolation-type | 10 (71) |
| **Gloves** |  |
| Single pair | 9 (64) |
| Double pair | 5 (36) |

Abbreviations: EGD, esophagogastroduodenoscopy; ERCP, endoscopic retrograde
cholangiopancreatography; EUS, endoscopic ultrasound.

**Figure 3.**
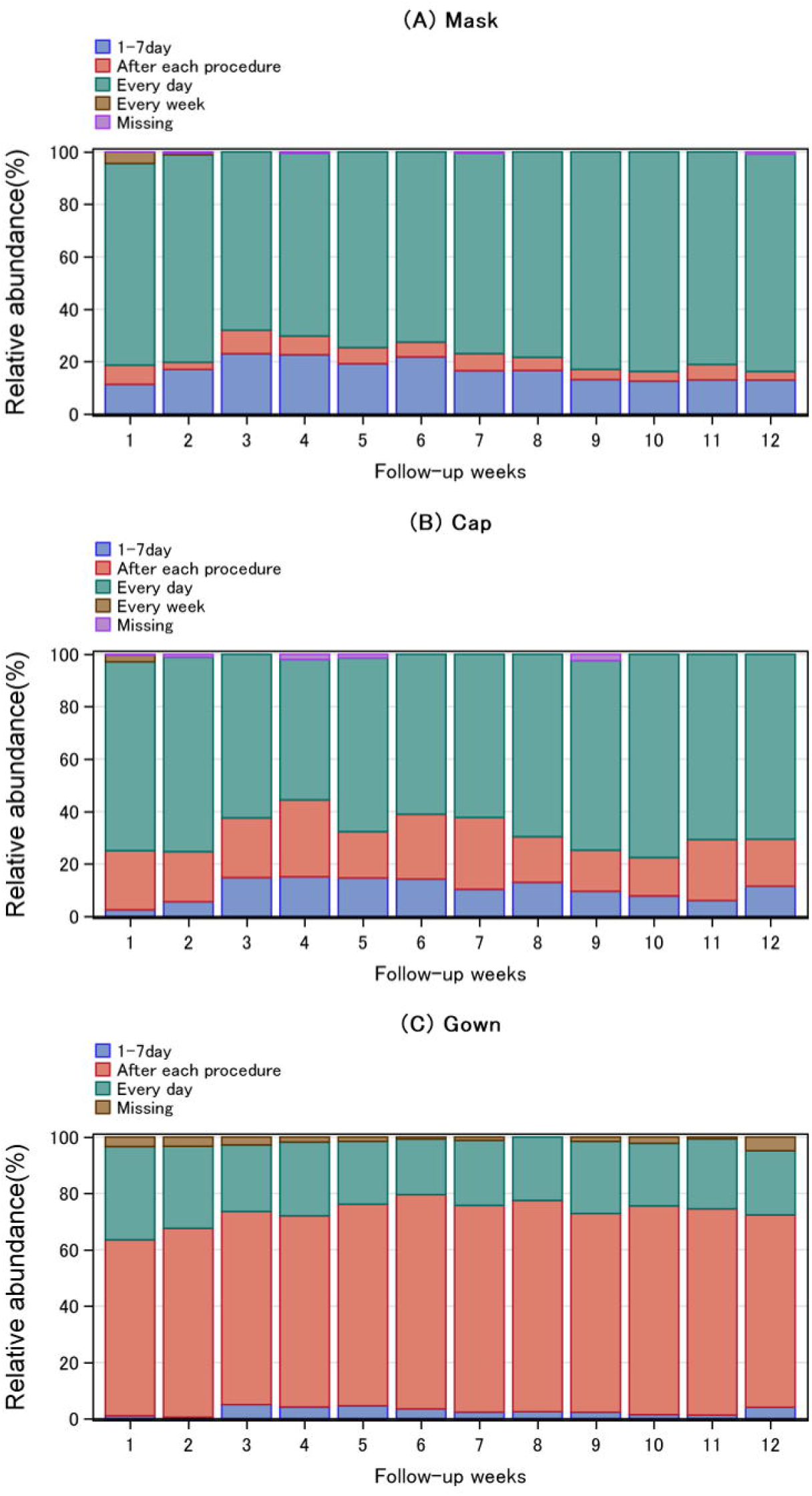
(A) COVID-19 incidence when treating COVID-19-positive patients (B) COVID-19 incidence when treating patients without confirmed COVID-19

The minimum PPE used was a surgical mask (21%), face shield but no goggles (14%), no cap (43%), short-sleeved vinyl-type gown (7%), and single pair of gloves (64%). The most common PPE used was a surgical mask plus N95 mask (36%) or N95 mask (36%), face shield but no goggles (50%), cap (57%), long-sleeved isolation-type gown (71%), and single pair of gloves (64%) (**Table 2**).

### COVID-19 incidence and the minimum and most common PPE used when treating patients without confirmed COVID-19

One participant who treated patients without confirmed COVID-19 developed COVID-19 over the mean follow-up period of 4.45 weeks (**Figure 3B**).

The minimum and most common PPE use with regard to masks was use of a surgical mask during all endoscopic procedures. The minimum PPE regarding face shields and goggles was no use of a face shield or goggles during all endoscopic procedures. The most common PPE use regarding face shields and goggles was no use of face shield but use of goggles. The minimum and most common PPE use regarding caps was use of a cap except for ESD. The minimum and most common PPE regarding gowns was no use of a gown and use of a long-sleeved isolation-type gown, respectively, during all endoscopic procedures. The minimum and most common PPE use regarding gloves was a single pair of gloves (**Table 3**).

**Table 3.**
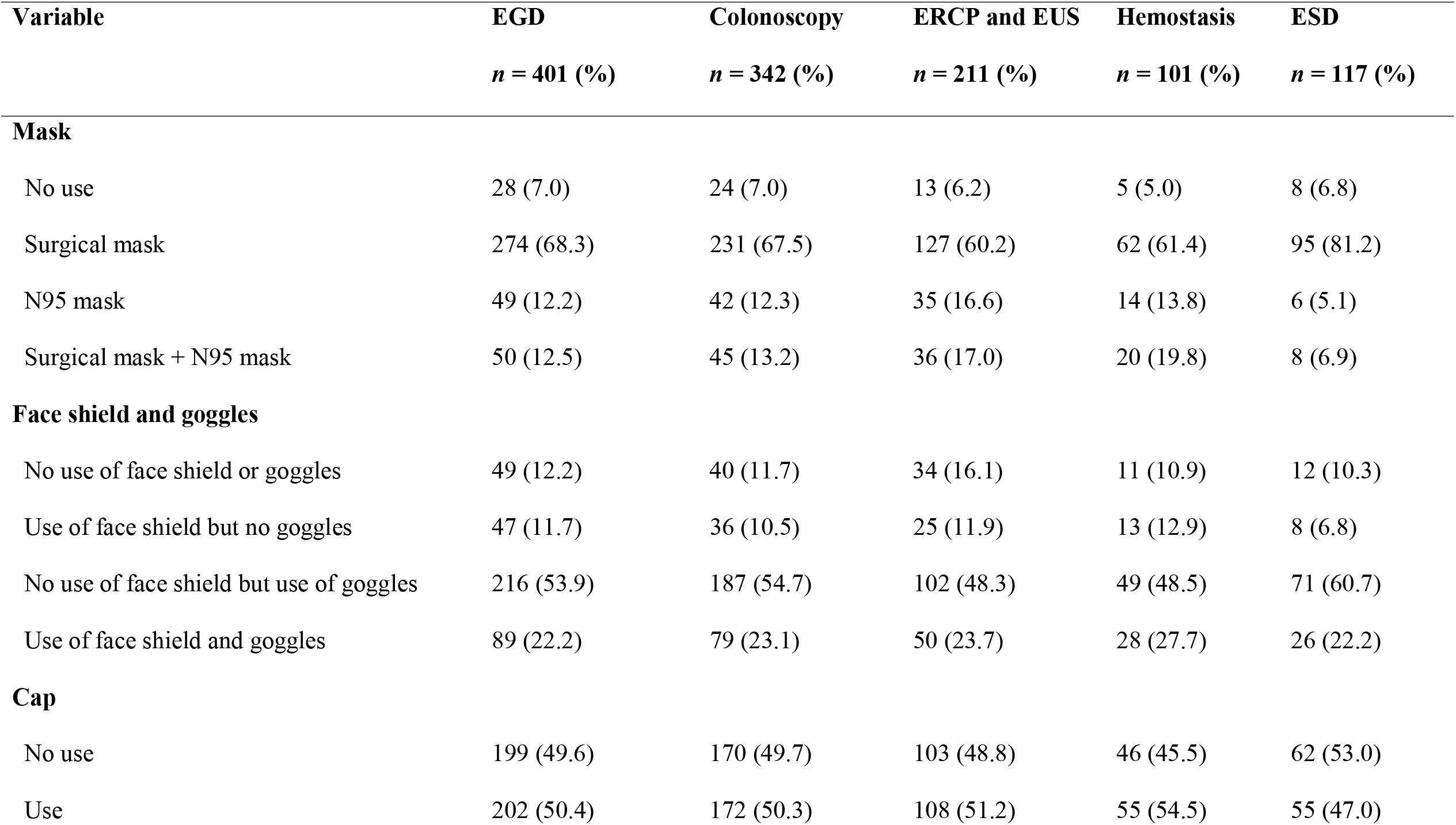
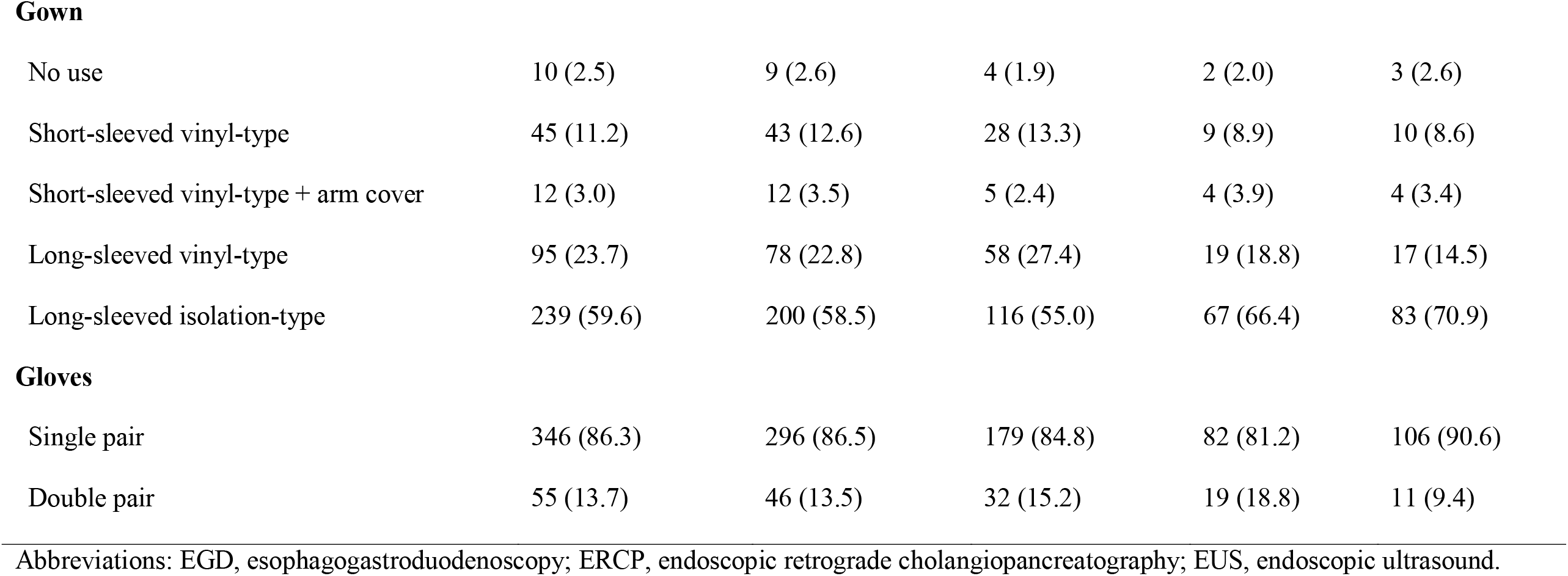
Endoscopic procedures and personal protective equipment used during procedures performed in patients without confirmed COVID-19.

| **Variable** | **EGD** | **Colonoscopy** | **ERCP and EUS** | **Hemostasis** | **ESD** |
| --- | --- | --- | --- | --- | --- |
|  | *n* = **401** (%) | *n* = **342** (%) | *n* = **211** (%) | *n* = **101** (%) | *n* = **117** (%) |
| **Mask** |  |  |  |  |  |
| No use | 28 (7.0) | 24 (7.0) | 13 (6.2) | 5 (5.0) | 8 (6.8) |
| Surgical mask | 274 (68.3) | 231 (67.5) | 127 (60.2) | 62 (61.4) | 95 (81.2) |
| N95 mask | 49 (12.2) | 42 (12.3) | 35 (16.6) | 14 (13.8) | 6 (5.1) |
| Surgical mask + N95 mask | 50 (12.5) | 45 (13.2) | 36 (17.0) | 20 (19.8) | 8 (6.9) |
| **Face shield and goggles** |  |  |  |  |  |
| No use of face shield or goggles | 49 (12.2) | 40 (11.7) | 34 (16.1) | 11 (10.9) | 12 (10.3) |
| Use of face shield but no goggles | 47 (11.7) | 36 (10.5) | 25 (11.9) | 13 (12.9) | 8 (6.8) |
| No use of face shield but use of goggles | 216 (53.9) | 187 (54.7) | 102 (48.3) | 49 (48.5) | 71 (60.7) |
| Use of face shield and goggles | 89 (22.2) | 79 (23.1) | 50 (23.7) | 28 (27.7) | 26 (22.2) |
| **Cap** |  |  |  |  |  |
| No use | 199 (49.6) | 170 (49.7) | 103 (48.8) | 46 (45.5) | 62 (53.0) |
| Use | 202 (50.4) | 172 (50.3) | 108 (51.2) | 55 (54.5) | 55 (47.0) |
| **Gown** |  |  |  |  |  |
| No use | 10 (2.5) | 9 (2.6) | 4 (1.9) | 2 (2.0) | 3 (2.6) |
| Short-sleeved vinyl-type | 45 (11.2) | 43 (12.6) | 28 (13.3) | 9 (8.9) | 10 (8.6) |
| Short-sleeved vinyl-type + arm cover | 12 (3.0) | 12 (3.5) | 5 (2.4) | 4 (3.9) | 4 (3.4) |
| Long-sleeved vinyl-type | 95 (23.7) | 78 (22.8) | 58 (27.4) | 19 (18.8) | 17 (14.5) |
| Long-sleeved isolation-type | 239 (59.6) | 200 (58.5) | 116 (55.0) | 67 (66.4) | 83 (70.9) |
| **Gloves** |  |  |  |  |  |
| Single pair | 346 (86.3) | 296 (86.5) | 179 (84.8) | 82 (81.2) | 106 (90.6) |
| Double pair | 55 (13.7) | 46 (13.5) | 32 (15.2) | 19 (18.8) | 11 (9.4) |

Abbreviations: EGD, esophagogastroduodenoscopy; ERCP, endoscopic retrograde cholangiopancreatography; EUS, endoscopic ultrasound.

### Change of PPE replacement frequency

Change of mask, cap, and gown replacement frequencies were same level at beginning and end of the study (**Figure 4**). Most common replacement frequency was every day in mask and cap, and each procedure in gown, respectively.

**Figure 4.**
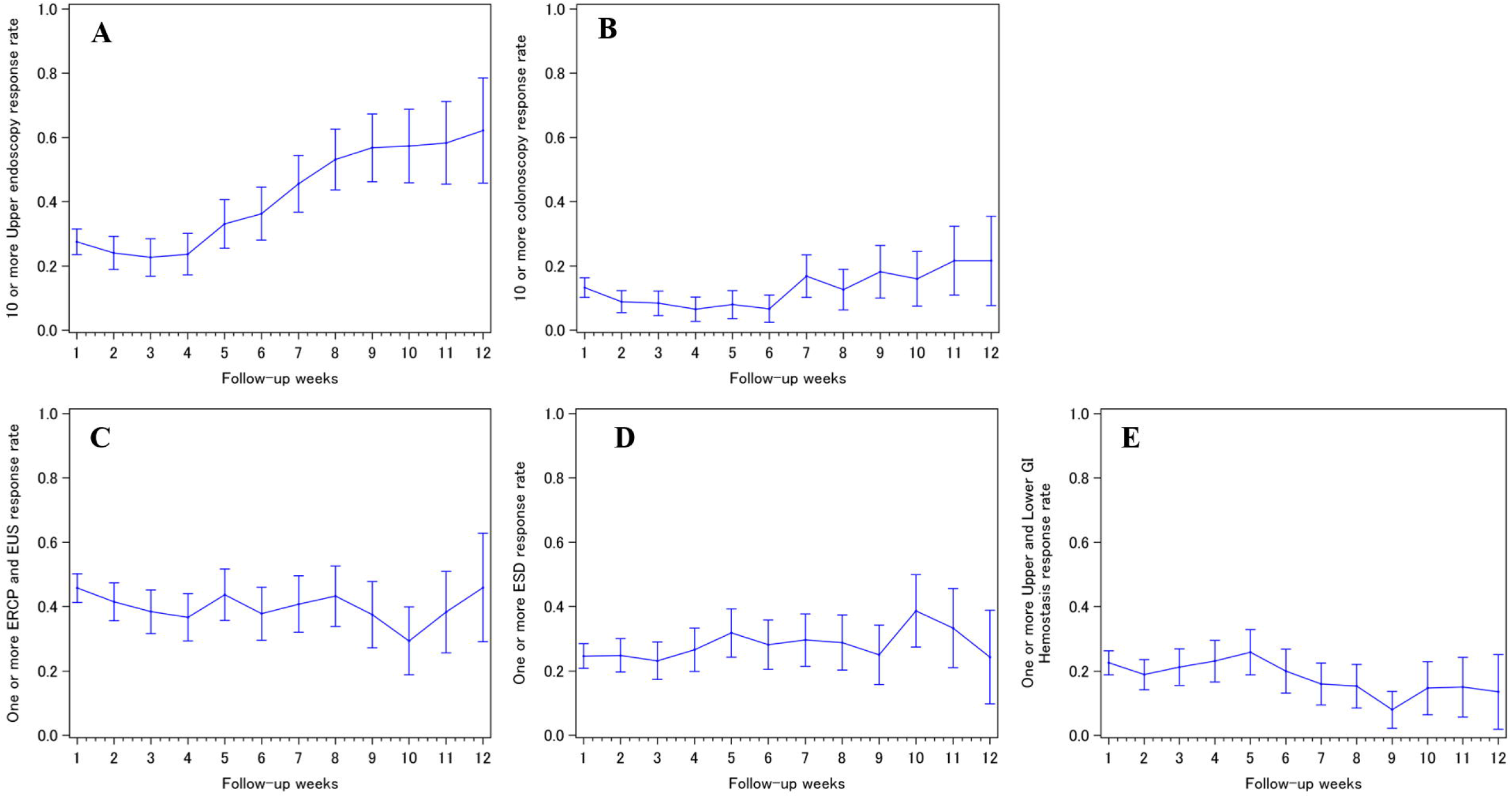
Change of personal protective equipment replacement frequency (A) Mask (B) Cap (C) Gown

## Discussion

In this international questionnaire survey, we found that the transmission risk of SARS-CoV-2 during endoscopic procedures was not high in appropriate PPE settings. In addition, our data suggest that the minimum PPE to prevent the development of COVID-19 was less than expected. In contrast to previous concerns, none of participant developed COVID-19 when treating COVID-19 positive patients and one participant developed COVID-19 when treating patients without confirmed COVID-19. There are two possible explanations for this discrepancy. First, most cases of nosocomial COVID-19 may occur when PPE is absent or insufficient in situations where healthcare professionals are unaware of the infection status of the patient. A recent European study also reported a low risk of COVID-19 transmission in endoscopy settings [14], which supported our findings. Another reason for the low incidence rate of COVID-19 in our study may be related to the procedure time and distance between the patient and healthcare professionals compared with other medical practices in the infectious ward. Most EGDs and colonoscopies take minutes to tens of minutes; upper and lower GI hemostasis and ERCP are completed within 1 hour, and even the longest ESD procedures are completed within a few hours. In addition, a certain distance is maintained between the patient and endoscopist, and no close contact with the patient is necessary.

The results of our survey identified the minimum PPE used during endoscopic procedures in patients diagnosed with COVID-19. With regard to masks, a surgical mask may have the potential to reduce COVID-19 transmission, it was consistent with previous observations [15]. The use of a face shield or goggles may help prevent transmission risk via the eyes. Therefore, we suggested the use of a surgical mask, or a face shield or goggles, any type of long-sleeved gown, and a single pair of gloves as the minimum PPE when performing EGD, colonoscopy, ERCP and EUS, upper and lower GI hemostasis, and upper GI ESD procedures in COVID-19-positive patients. However, further studies to collect data from more endoscopic procedures conducted in patients with a COVID-19 over longer-term observation periods are needed to elucidate the appropriate PPE to prevent the development of COVID-19 related to endoscopic procedures.

In contrast to many health care providers concerns, replacement frequencies of mask, cap, and gown were maintained at the similar levels throughout the study period (Figure 4). The most common timing to replacement was “every day” in mask and cap, and “after each procedure” in gown, respectively.

This was the first longitudinal international survey to evaluate the incidence of nosocomial COVID-19 among healthcare professionals involved in endoscopic procedures. Our survey included participant from all continents except Antarctica and included both areas with high and low rates of SARS-CoV-2 infection. In addition, we used a solid questionnaire survey system to collect high-quality questionnaire data. Repeat questionnaire surveys will enable us to perform survival analyses of patients with COVID-19. However, our study had some limitations. First, the primary outcome data were collected using self-reported questionnaires only, and we did not perform additional surveys, such as collection of PCR or computed tomography data, to confirm the events to protect the privacy of the participants. In addition, we may have missed some COVID-19 events because some participants developed severe pneumonia and did not respond to the survey during the follow-up period. However, we invited participants using a personal private network among academic experts, and we are continuously collecting data. Therefore, we believe the data to be both reliable and of high quality. Second, differential diagnosis of community-acquired and endoscopy-related SARS-CoV-2 infections may be difficult. Third, our primary outcome definition was COVID-19 development, not COVID-19 pneumonia, or other symptoms development.

In conclusion, the risk of COVID-19 transmission during endoscopic procedures, including EGD, colonoscopy, ERCP and EUS, upper and lower GI hemostasis, and upper GI ESD, was low in PPE settings.

## Data Availability

Raw data usage is limited to principal investigator team.
Summarized data usage is open on the web.

https://sites.google.com/g.ecc.u-tokyo.ac.jp/anticovid19

## Acknowledgments

We thank all the participants and their families.

## Statement of Ethics

The study was approved by the Ethics Committee of the University of Tokyo Hospital (No. 2020046NI) and conducted ethically in accordance with the World Medical Association Declaration of Helsinki. Informed consents were obtained by web survey.

## Author Contributions

All authors contributed to the design of the study. T. Kawahara performed the data management and statistical analysis. R. Niikura drafted the manuscript. All authors have read and approved the final manuscript.

