## Supplementary Table 1 for "International observational survey of the effectiveness of personal protective equipment during endoscopic procedures performed in patients with COVID-19"

**Supplementary Table 1. Questionnaire**

1. What is your age?

2. What is your sex?

Response: female/male/no response

3. Where do you work in endoscopy?

Response: Asia/Europe/North America/South America/Oceania/Africa

4. What is your role in your unit?

Response: Doctor or endoscopist/nurse/laboratory technician/cleaner

5. Number of years of experience?

Response: < 5 years/5 to < 10 years/≥ 10 years

6. Have you been diagnosed with COVID-19?

Response: No/Yes

7. Did you perform endoscopic examinations in COVID-19 patients?

Response: No/Yes

8. What endoscopic examinations did you perform in non-COVID-19 patients? * Multiple responses allowed

Response: Esophagogastroduodenoscopy/colonoscopy/ERCP and EUS/upper GI hemostasis/lower GI hemostasis/upper GI ESD/lower GI ESD

9. Which type of mask did you use? * Multiple responses allowed

Response: Surgical mask/N95 (United States NIOSH-42CFR84)/FFP2 (Europe EN 149-2001)/KN95 (China GB2626-2006)/P2 (Australia/New Zealand AS/NZA 1716:2012)/Korea 1st class (Korea KMOEL - 2017-64)/DS (Japan JMHLW-Notification 214, 2018)/Face shield mask

10. How often did you change the mask/shield?

Response: After each procedure/every day/after more than 1 day but less than 1 week

11. Did you use a cap?

Response: No/Yes

12. How often did you replace the cap?

Response: After each procedure/every day/after more than 1 day but less than 1 week

13. Did you use goggles?

Response: No/Yes

14. What type of gown did you use?

Response: No gown/short-sleeved vinyl-type gown + arm cover/short-sleeved vinyl-type gown/long-sleeved vinyl-type gown/long-sleeved isolation-type gown

15. How often did you replace the gown?

Response: After each procedure/every day/after more than 1 day but less than 1 week

16. Which style of gloves do you currently use?

Response: No gloves/single pair/double pair
